# Biologically contained Ebola virus enables standardised neutralisation testing for preclinical and clinical immunogenicity assessment

**DOI:** 10.64898/2026.02.06.26345727

**Authors:** Jens Verlinden, Ola Diebold, Dung Nguyen, Joseph Akoi-Boré, Bert Vanmechelen, Stephen M. Laidlaw, Piet Maes, Miles W. Carroll

## Abstract

**Background:** Neutralising antibody titres are widely used as key immunogenicity endpoints in Ebola virus (EBOV) vaccine and monoclonal antibody clinical trials. However, direct comparison of results across studies remains challenging due to the use of heterogeneous neutralisation platforms, ranging from pseudotyped viruses to live EBOV assays. These limitations restrict assay standardisation, validation, scalability, and compliance with good clinical laboratory practice (GCLP), particularly in outbreak-prone and resource-limited settings. There is an unmet need for neutralisation assays that combine biological authenticity with clinical-trial compatibility.

**Methods:** We developed and optimised a fluorescence-based microneutralisation assay using a biologically contained EBOV lacking the essential VP30 gene (EBOVΔVP30), enabling multi-cycle viral replication under containment level 2 conditions. Using a defined panel of serum samples from Ebola virus disease survivors and EBOV-negative controls, we benchmarked EBOVΔVP30 neutralisation titres against previously generated data obtained with wild-type EBOV and pseudotyped virus platforms. Assay performance was evaluated in terms of sensitivity, reproducibility, discrimination between positive and negative samples, and correlation with live virus neutralisation. Calibration was performed using the WHO International Standard for anti-EBOV immunoglobulin.

**Findings:** The EBOVΔVP30 microneutralisation assay robustly distinguished EBOV survivor sera from negative controls (p < 0·0001) and demonstrated a strong correlation with live EBOV neutralisation titres (Spearman ρ = 0·8725). This correlation exceeded that observed for HIV-1-based pseudotyped assays and for the vesicular stomatitis virus-based platforms. The fluorescence-based read-out showed comparable sensitivity to conventional immunostaining, supporting its suitability for high-throughput and standardised implementation. Importantly, assay conditions were compatible with BSL-2 laboratories and GCLP-aligned workflows.

**Interpretation:** Biologically contained EBOVΔVP30 provides a clinically relevant and scalable alternative to existing neutralisation platforms, bridging the gap between pseudotyped assays and wild-type virus testing. By improving biological relevance while maintaining accessibility and standardisation, this assay has the potential to enhance comparability of immunogenicity data across EBOV vaccine and therapeutic antibody (pre-)clinical trials, aligning with global outbreak preparedness and trial harmonisation objectives.

**Funding:** Stated in acknowledgement section of manuscript.

**Research in context:** *Evidence before the study:* Before starting this study, we reviewed published work on how neutralising antibodies against Ebola virus are measured in vaccine and monoclonal antibody research. We searched PubMed, Web of Science, and reference lists of key review papers for studies published up to mid-2025, without restricting by language. Search terms included “Ebola virus”, “neutralising antibodies”, “neutralisation assay”, “pseudovirus”, “live virus”, and “clinical trials”. We focused on studies describing neutralisation tests using wild-type Ebola virus as well as commonly used pseudotyped virus systems. From this body of evidence, neutralisation assays using wild-type Ebola virus are considered the most biologically relevant but can only be performed in biosafety level 4 laboratories. This limits their availability, scalability, and use in clinical trials. Pseudotyped virus assays can be performed under lower biosafety conditions and are widely used, but multiple studies have reported variable performance and inconsistent agreement with live virus results. Although biologically contained Ebola viruses have been developed and used in laboratory research, their application as neutralisation assays and their direct comparison with both live virus and pseudotyped systems using the same human serum samples had not been systematically studied. As a result, it remained unclear whether such systems could support reliable immunogenicity assessment in clinical trials.

*Added value of this study:* This study shows that a biologically contained Ebola virus lacking the VP30 gene can be used to measure neutralising antibodies in a robust and scalable way under biosafety level 2 conditions. By directly comparing this system with wild-type Ebola virus and widely used pseudotyped assays using the same set of human serum samples, we demonstrate that neutralisation results obtained with the biologically contained virus closely align with those of the wild-type virus reference assay. The assay reliably distinguishes samples from Ebola survivors and uninfected individuals and can be read using different detection methods, making it compatible with GCLP-aligned workflows and suitable for further qualification and validation in support of clinical development. This work provides clear evidence that biologically contained Ebola virus can combine biological relevance with practical usability.

*Implications of all the available evidence:* Together with existing evidence, our findings indicate that biologically contained Ebola virus offers a valuable new option for measuring neutralising antibodies in vaccine and monoclonal antibody clinical trials. By reducing reliance on high-containment laboratories while preserving key features of authentic virus infection, this approach can improve the consistency and comparability of immunogenicity data across studies and sites. Broader use of such assays could support better decision-making during clinical development and strengthen outbreak preparedness. More generally, this work highlights how biologically contained viruses can help advance research licensure of medical countermeasures for high-consequence pathogens in ways that are directly relevant to human health.

## Introduction

Ebola virus (EBOV) remains a major global public health threat, causing recurrent outbreaks of viral haemorrhagic fever with high case fatality rates (up to 90%)^1^. Despite significant advances in outbreak response, including the deployment of effective vaccines and monoclonal antibody therapies, Ebola virus disease (EVD) continues to pose substantial challenges, particularly in resource-limited and outbreak-prone regions^2,3^. Sustained preparedness efforts therefore rely not only on the availability of medical countermeasures but also on robust tools to evaluate immune responses induced by infection, vaccination, or therapeutic intervention.

Neutralising antibody responses are widely regarded as a key functional correlate of protective immunity against EBOV and are routinely used as primary or secondary immunogenicity endpoints in vaccine and therapeutic antibody clinical trials^4–7^. Accurate measurement of neutralising activity is therefore critical for the interpretation of clinical trial outcomes, comparison of vaccine platforms, and assessment of immune durability in survivors and vaccinated populations. In addition, reliable neutralisation assays are essential for sero-epidemiological studies aimed at defining population-level exposure and immunity, which inform outbreak risk assessment and preparedness strategies^8^.

A broad range of serological assays is available to assess EBOV-specific immune responses. Binding assays such as enzyme-linked immunosorbent assays (ELISA), multiplex bead-based platforms, immunofluorescence assays, and western blotting are widely used due to their scalability and relative simplicity^9–12^. While these approaches provide valuable information on antibody presence and specificity, they do not directly assess functional virus neutralisation. Label-free technologies such as surface plasmon resonance and bio-layer interferometry offer detailed insights into antibody–antigen binding kinetics but are less commonly applied in clinical trial settings due to their technical complexity, cost, and limited correlation with in vivo protection^13,14^. As a result, neutralisation assays remain the gold standard for evaluating functional humoral immunity in EBOV research and clinical development^15^.

The accepted reference for measuring EBOV neutralising antibodies is the plaque reduction neutralisation test (PRNT) or focus reduction neutralisation test (FRNT) using wild-type live virus^15–17^. These assays provide high biological relevance but require biosafety level 4 (BSL-4) containment, are labour-intensive, and are poorly compatible with high-throughput workflows or good clinical laboratory practice (GCLP). Consequently, their use is restricted to a limited number of specialised facilities, hindering assay standardisation and cross-study comparability in multicentre clinical trials.

To overcome these constraints, pseudotyped virus neutralisation assays have been widely adopted. These systems employ non-pathogenic viral backbones, such as vesicular stomatitis virus (VSV) or human immunodeficiency virus type 1 (HIV-1), engineered to express EBOV glycoprotein (GP), enabling EBOV entry to be studied under BSL-2 conditions^18,19^. Pseudotyped assays have proven valuable for high-throughput screening and have been used extensively in EBOV vaccine development, as well as for other viral pathogens^20–22^. However, accumulating evidence indicates that pseudotyped systems may not fully recapitulate the structural, genetic, and replicative complexity of wild-type EBOV. Differences in viral morphology, GP processing, co-transcriptional editing, and particle assembly can influence neutralisation sensitivity and antibody ranking^19,23–25^. Importantly, these assay-dependent differences introduce variability that may complicate the interpretation and comparison of immunogenicity data across clinical trials.

Biologically contained EBOV systems offer a promising alternative that addresses many of these limitations. VP30 is an essential transcription factor required for productive EBOV replication. By deleting this gene, viral replication becomes restricted to genetically engineered cell lines that supply VP30 in trans, enabling multi-cycle replication under BSL-2 conditions.^26–28^. These systems preserve wild-type EBOV genome organisation, replication dynamics, and virion morphology while maintaining an improved biosafety profile. Although biologically contained EBOV has been used in antiviral screening and mechanistic studies, its application in antibody neutralisation testing has remained limited, and direct benchmarking against both live virus and pseudotyped assays using identical clinical serum panels has not been systematically performed^29^.

In this study, we aimed to address this gap by developing and optimising a fluorescence-based microneutralisation assay using biologically contained EBOV lacking VP30 (EBOVΔVP30). Using a defined panel of serum samples from EVD survivors and EBOV-negative individuals previously characterised by live virus and pseudotyped neutralisation assays^19^, we performed a direct, pairwise comparison across platforms. By benchmarking assay sensitivity, discrimination, and correlation with wild-type EBOV neutralisation, we sought to evaluate the suitability of EBOVΔVP30 as a clinically relevant and scalable neutralisation platform. Our findings have direct implications for the standardisation and comparability of immunogenicity assessments in EBOV vaccine and monoclonal antibody clinical trials and support the broader implementation of biologically contained systems in outbreak preparedness and translational research.

## Materials and methods

### Cell lines

For this study, African Green monkey kidney cells (Vero E6; Vero C1008, ATCC) expressing EBOV (strain Mayinga) VP30 were used. Generation of this modified cell line was described previously^28^. Cells were cultivated in Dulbecco’s Modified Eagle Medium (DMEM; Thermo Fisher Scientific), supplemented with 10% Foetal Bovine Serum (FBS; Biowest, Nuaillé, France), 200 mM L-Glutamine (Thermo Fisher Scientific) and 1% sodium bicarbonate (Thermo Fisher Scientific). As standard antibiotics, 1% Penicillin-streptomycin (Thermo Fisher Scientific), 0·2% Fungizone (Thermo Fisher Scientific) and 0·02% Gentamicin (Thermo Fisher Scientific) were used. Additionally, Blasticidin S (10 mg/mL) was added to culture medium to prevent the loss of integrated VP30 constructs after extensive passaging. Whole-genome sequencing after ten serial passages confirmed genetic stability of EBOVΔVP30, with no evidence of VP30 reversion (supplementary Figure 1). Cells were passaged and maintained at 37 °C with 5% CO_2_.

### Viruses

Biologically contained EBOV (EBOVΔVP30) was obtained as previously described^28^. Briefly, the virus was rescued by transfecting the VP30-expressing Huh-7 cells with an EBOV antigenome and essential support plasmids encoding the nucleocapsid proteins needed for the first transcription and encapsidation round. Supernatants from cells showing eGFP expression were sequentially passaged on VeroE6 cells expressing EBOV VP30 to amplify the virus stock. Virus was harvested through centrifugation for 15 minutes at 4500xg and stored at -80 °C. Virus stocks were routinely sequenced prior to use in the assays to rule out potential mutations in relevant regions.

### WHO Reference Standard 15/220

The WHO International Standard for anti-EBOV immunoglobulin (National Institute for Biological Standards and Control; NIBSC; code 15/220) was used to calibrate the neutralisation assay^30^. This polyclonal plasma preparation serves as a globally accepted reference material for assay standardization and inter-laboratory comparison of EBOV neutralising antibody responses. The standard was reconstituted and used in accordance with the manufacturer’s instructions provided by the NIBSC. Therefore, 0·5ml of Dulbecco’s phosphate-buffered saline (DPBS, Gibco) was added to the purified material.

### Human Serum samples

The serum samples, as described and tested by Steeds *et al*., were obtained from a pre-existing biobank generated from prior studies on Ebola virus survivors from the forested region of Guinea^19^. Prior to use in our neutralization assay, serum samples were heat inactivated for 30 minutes at 56 °C.

### Focus-forming assay

To estimate the concentration of our virus stocks, we initially performed a focus-forming assay in 6-well plates. VeroE6.EBOV.VP30 cells were seeded at a density of 700,000 cells per well. The following day, the culture medium was removed and 200 µL of serially diluted virus stock (ranging from 10^−1^ to 10^−5^) was added in duplicate. Plates were incubated at 37 °C and 5% CO_2_ for one hour, with gentle swirling every 10-15 minutes to ensure even virus distribution. After incubation, 3 mL of overlay medium containing agarose was added to each well. This overlay was prepared by mixing autoclaved 1·76% SeaKem ME agarose 1:1 with DMEM supplemented with 20% FBS, 200 mM L-glutamine, 1% penicillin-streptomycin, 2% non-essential amino acids, 1% gentamicin, and 0·2% Fungizone. Once cooled to 37 °C, the mixture was applied to the wells and allowed to solidify. Plates were then incubated for three days, after which fluorescent foci (eGFP-positive foci) were manually counted.

To determine the optimal virus input for our microneutralization assay, we further conducted a focus-forming assay in 96-well format to estimate the amount of virus stock required to achieve approximately 100 focus-forming units (FFU) per well, a commonly accepted input for PRNT-based assays^6,31^. Based on the results of the 6-well focus-forming assay, a range of infectious doses was tested around the estimated concentration. VeroE6-EBOV-VP30 cells were seeded at 20,000 cells per well and incubated overnight. The following day, a 10-fold 1:2 serial dilution of the virus stock was prepared and added to the wells in six replicates. Plates were incubated at 37 °C with 5% CO_2_, and foci were manually counted at 24 and 45 hours post-infection (hpi).

### Micro-neutralization assay

For the microneutralisation assay, VeroE6.EBOV.VP30 cells were seeded at a density of 20,000 cells per well in 96-well plates using DMEM supplemented with 10% foetal bovine serum (FBS) and incubated overnight at 37 °C with 5% CO_2_. On the following day, an 8-fold, 1:2 serial dilution of human serum samples was prepared in DMEM (+10% FBS) in a separate 96-well plate, starting at a 1:10 dilution in a total volume of 20 µL per well. Each sample was tested in duplicate in two independent experiments, resulting in four replicates per sample. Simultaneously, virus stock was diluted in DMEM (+10% FBS) to a concentration of 6·67 × 10^3^ FFU/mL, and 20 µL of this virus suspension was added to each well containing the serum. The virus–serum mixtures were incubated at 37 °C with 5% CO_2_ for 1 hour to allow binding between the virus and neutralizing antibodies in the sera. Culture medium was then removed from the pre-seeded cell plates and 30 µL of the virus–serum mixture was transferred to the corresponding wells, resulting in a final input of 100 FFU per well. Each assay plate included six negative control wells (medium only) and six virus control wells (virus without serum). Plates were incubated for 1 hour at 37 °C with 5% CO_2_, with gentle swirling every 15 minutes to ensure even virus distribution. Following adsorption, 100 µL of DMEM containing 10% FBS was added to each well. Plates were incubated for 45 hours at 37 °C with 5% CO_2_. The inoculum was removed from all wells whereafter 100 µL DPBS was added to reduce background signal. As no overlay medium or fixation was applied, plates were kept at room temperature in case read-out was postponed, preventing the development of secondary foci.

### Plate read-out

For the eGFP-read-out, plates were imaged and analysed using an ImmunoSpot S6 Analyzer (Cellular Technology Limited, CTL) equipped for eGFP fluorescence detection. Foci, appearing as discrete eGFP-positive foci, were automatically counted using the ImmunoSpot Fluoro-X™ Suite software version 7.0.30.4. The counting parameters were optimized and pre-defined on a selection of typical wells representative of the response(s) seen in the plate(s) (including both negative control and positive wells). Subsequently, auto count was performed on all plates using the pre-set template followed by a quality control phase during which a variety of artefacts, such as overlapping signals or damaged membranes were further refined.

### Immunostaining

In preparation for immunostaining, DPBS was removed and cells were fixed with 4% v/v paraformaldehyde (Alfa Aesar) (100 µL/well) for 30 min at room temperature. Cells were then permeabilised with Triton-X 100 (1% in PBS) and stained for EBOV GP using a human monoclonal antibody (#141) (1:5000) (gifted by dr. Pramila Rijal and Dr Francesca Donnellan^32^). Bound antibody was detected following 1 hour incubation with goat anti-human IgG horseradish peroxidase conjugate (Sigma-Aldrich) (1:5000). Subsequently, plates were washed with PBS/Tween 0·1% and TrueBlue Peroxidase substrate (Insight Biotechnology) was added to visualise the foci. Plates were then imaged and foci counted using an ELISPOT reader.

### Statistical analysis

All analyses were performed using R studio v4.2.2 and GraphPad Prism v10.6.1. To determine the neutralising capacity of the WHO reference standard 15/220 and human serum samples, a four-parameter logistic (4PL) non-linear regression model was applied to normalized data. Normalisation was performed relative to virus-only control wells, and results were expressed as focus forming units (FFU) in percentages (%), allowing for consistent comparisons across plates. Spearman rank correlation was used to assess the relationship between EBOVΔVP30 and both pseudotyped and wild-type live virus assays. A two-tailed Mann-Whitney U test was conducted to statistically evaluate differences in neutralisation titres between negative samples and samples from EBOV survivors and a Wilcoxon signed-rank test was performed to compare assay sensitivity between eGFP- and immunostaining read-out. For paired comparison of eGFP- and immunostaining-derived NT_50_ values, a Wilcoxon signed-rank test was used to assess systematic differences between read-out methods.

### Ethics statement

All methods were carried out in accordance with the relevant guidelines and regulations under ethical approval from the Guinean Research Ethics Committee; No. 33/CNERS/15.

## Results

### EBOVΔVP30 microneutralisation discriminates clinically relevant antibody responses

To evaluate the clinical relevance of the EBOVΔVP30-based microneutralisation assay, we applied the optimised conditions to a defined panel of 40 human serum samples previously characterised in EBOV neutralisation studies^19^. This panel comprised 30 sera from EVD survivors with a broad range of neutralising antibody titres and 10 sera from confirmed EBOV-negative individuals.

Neutralising titres (NT_50_) were calculated using a four-parameter logistic regression model based on four replicates per sample. A conservative assay threshold was defined based on the highest serum concentration tested (1:20), corresponding to the lowest limit of detection. Using this approach, the EBOVΔVP30 assay robustly distinguished EVD survivor sera from negative controls (p = 3·76 × 10^-6^, Mann-Whitney U test, Cohen’s d = 0·83) (Figure 1).

**Figure 1.**
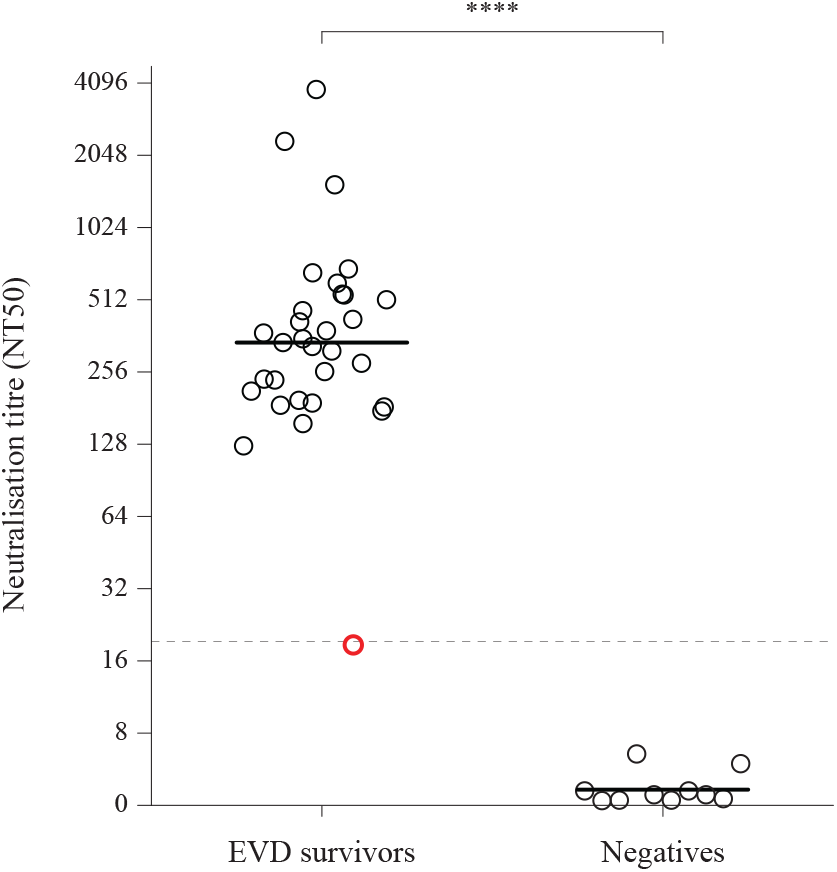
EBOVΔVP30 microneutralisation discriminates clinically characterised EBOV survivor sera from negative controls. Neutralising antibody titres (NT50) determined using the EBOVΔVP30 microneutralisation assay in serum samples from Ebola virus disease survivors (n = 30) and EBOV-negative individuals (n = 10). Each circle represents an individual serum sample. Horizontal bars indicate the median for each group. The dashed line indicates the assay threshold (NT50 = 20). Red circle denotes a sample showing a discordant result. Neutralising titres were calculated by non-linear regression and are displayed on a log_2_ scale. Statistical significance between groups was assessed using a two-tailed Mann-Whitney U test (**** p < 0·0001). Data represent the mean of three technical duplicates derived from independent experiments.

Only one survivor serum (G022) failed to exceed the assay threshold. Retrospective analysis of live virus data indicated that this sample previously displayed a borderline neutralisation titre below the accepted geometric mean threshold, suggesting that the EBOVΔVP30 result reflects a genuinely low neutralising response rather than assay insensitivity. Overall, these findings demonstrate that EBOVΔVP30 reliably discriminates clinically relevant neutralising antibody responses in human sera.

### EBOVΔVP30 neutralisation titres strongly correlate with wild-type EBOV

To assess the biological relevance of the EBOVΔVP30 platform, neutralisation titres obtained in this study were directly compared with previously generated data from wild-type EBOV and pseudotyped virus assays using the same serum panel^19^. This enabled a robust, pairwise benchmarking across platforms.

A strong correlation was observed between EBOVΔVP30 and live EBOV neutralisation titres (Spearman ρ = 0·8725, p = 2·22 × 10^-13^, and Log_10_-transformed Pearson correlation r = 0·843, p = 8·15 × 10^-8^) (Figure 2A), indicating close alignment with the reference assay performed under BSL-4 conditions. In comparison, EBOVΔVP30 titres showed a lower correlation with vesicular stomatitis virus (VSV)-based pseudotyped assays (ρ = 0·741, p < 0·001) (Figure 2C) and HIV-1-based pseudotyped assays (ρ = 0·446, p = 0·014) (Figure 2B).

**Figure 2.**
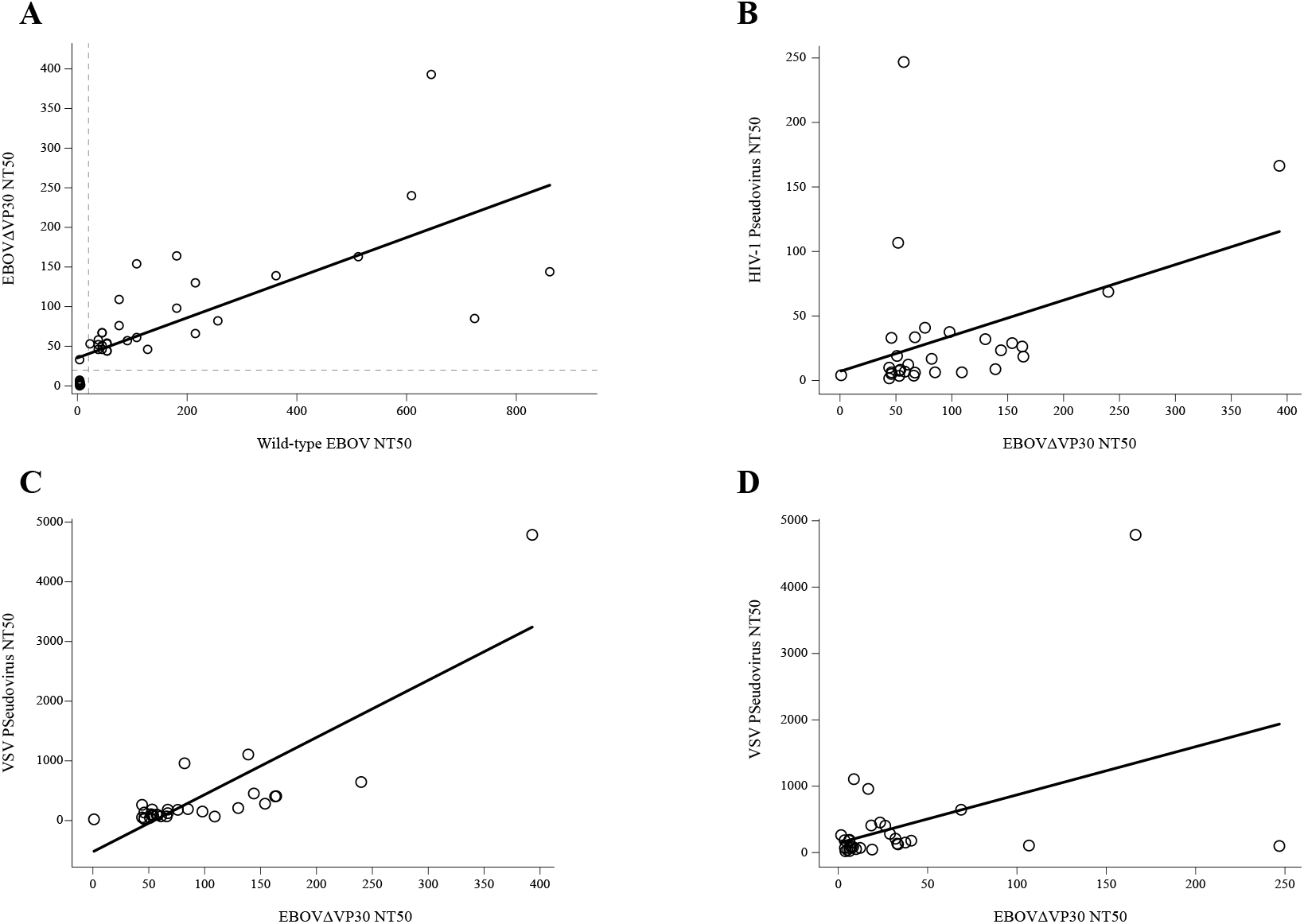
Correlation analysis between neutralization assays. Scatter plots showing correlations between the (A) wild-type EBOV and EBOVΔVP30 assays (ρ = 0·8725, p = 2·22 × 10-13), (B) EBOVΔVP30 and HIV-1 pseudovirus assays (ρ = 0·446, p = 0·014), (C) EBOVΔVP30 and VSV pseudovirus assays (ρ = 0·741, p < 0·001), and (D) HIV-1 pseudovirus and VSV pseudovirus assays (ρ = 0·369, p = 0·017) in serum samples from EBOV survivors (n = 40). (ρ) Spearman’s rank correlation coefficient. All p-values < 0·05 were considered statistically significant.

These results demonstrate that EBOVΔVP30 closely reflects wild-type virus neutralisation and performs at least comparably to commonly used pseudotyped platforms, while outperforming HIV-1-based systems.

### Optimisation of assay conditions enables robust and standardised implementation

To ensure reproducibility and suitability for standardised application, key assay parameters were systematically optimised, including read-out timing, viral input, and the use of overlay medium.

Initial experiments assessed the earliest time point at which eGFP-positive foci could be reliably quantified. At 24 hpi, fluorescence signals were weak and variable, resulting in poor dose– response characteristics (Figure 3A). In contrast, read-out at 45 hpi yielded strong and uniform fluorescence with well-defined foci and a reproducible sigmoidal neutralisation curve using the WHO International Standard for anti-EBOV immunoglobulin (Figure 3A). Accordingly, 45 hpi was selected as the optimal read-out time for subsequent experiments.

**Figure 3.**
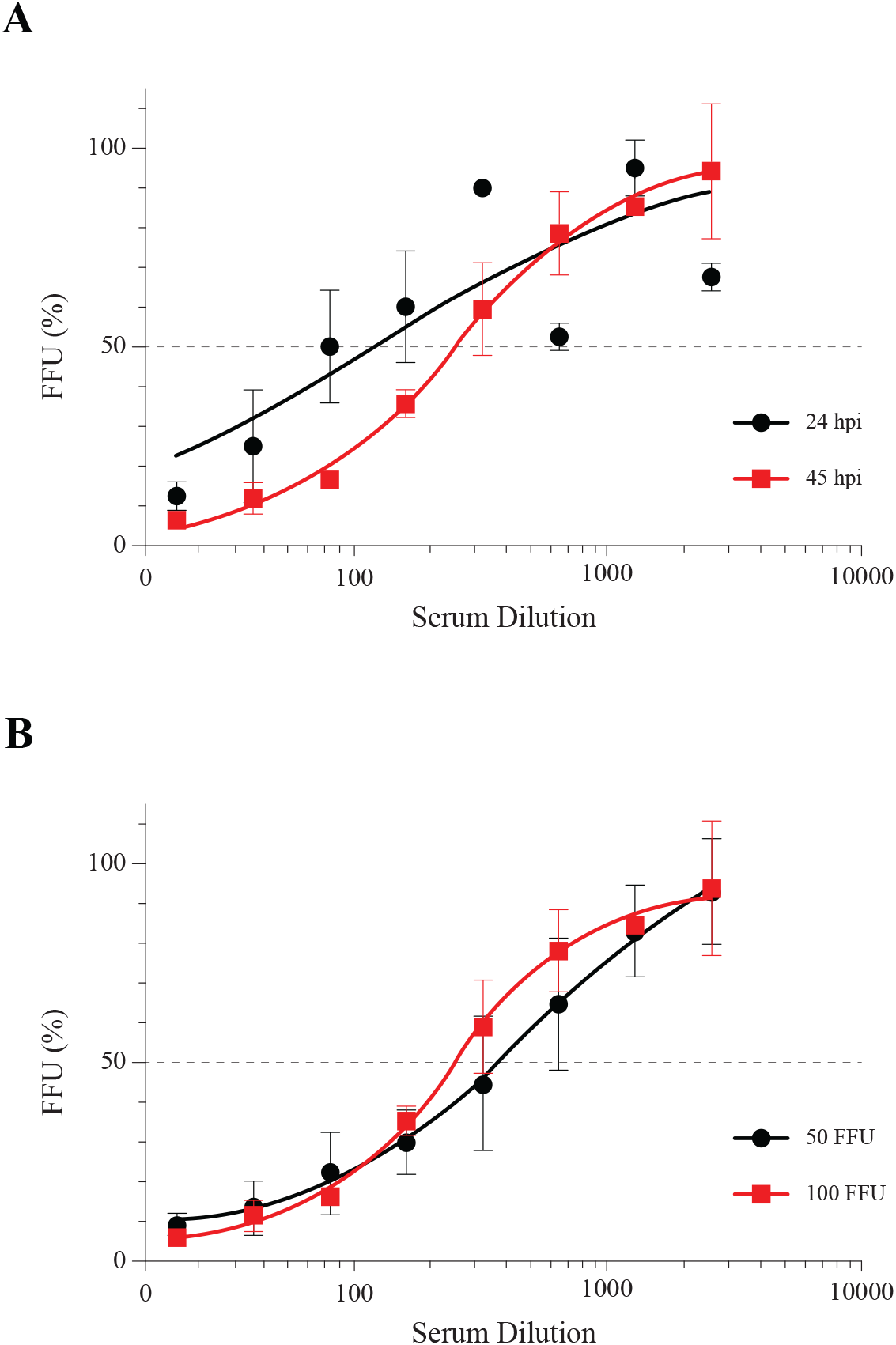
Optimisation of assay conditions supporting standardised EBOVΔVP30 microneutralisation. Optimisation of infectious dose and read-out timing for the EBOVΔVP30 microneutralisation assay using the WHO International Standard for anti-EBOV immunoglobulin (15/220). *(A)* Neutralisation curves showing normalised EBOVΔVP30 focus-forming units (FFU) at 24 hours post-infection (hpi) and 45 hpi, demonstrating improved signal robustness and reproducibility at the later time point. *(B)* Comparison of neutralisation curves obtained using infectious doses of 50 or 100 FFU per well, supporting the selection of 100 FFU to align with established reference standards and enable assay standardisation. Data represent mean ± SD of three replicates.

We next evaluated the impact of viral input on assay sensitivity. Both 50 and 100 FFU per well produced clear dose–response curves (Figure 3B). However, the lower viral input resulted in higher calculated NT_50_ values that exceeded those reported in international standardisation studies. To maintain alignment with existing reference data and accepted PRNT practices, 100 FFU per well was selected as the standard assay input.

The use of a carboxymethylcellulose overlay was also assessed but introduced increased technical variability between wells and plates (data not shown). Consequently, the overlay was omitted in favour of a simplified protocol, with plates transferred to room temperature after 45 hpi to limit secondary viral spread.

### Flexible read-out strategies support scalable deployment

To further validate the robustness and versatility of the EBOVΔVP30 microneutralisation assay, neutralisation titres derived from eGFP-based fluorescence detection were compared with those obtained using conventional immunostaining on the same assay plates.

A strong correlation was observed between NT_50_ values generated by eGFP fluorescence and immunostaining read-outs (Spearman ρ = 0·868, p < 0·0001) (Figure 4A).

**Figure 4.**
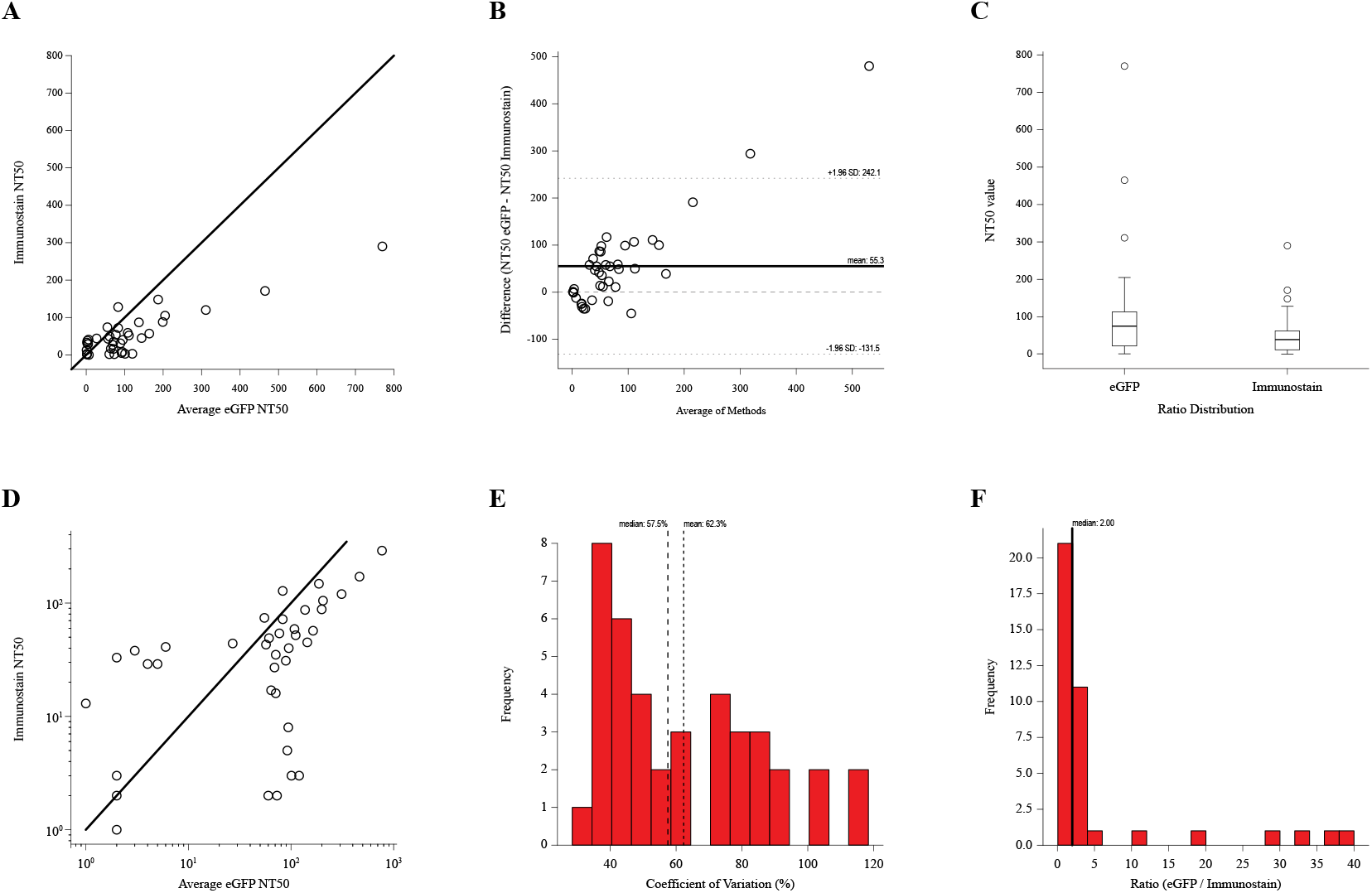
Fluorescence-based and immunostaining read-outs show strong correlation with a systematic difference in absolute neutralisation titres. *(A) Correlation Plot*. Scatter plot showing the relationship between average eGFP NT50 values and immunostaining NT50 values for 40 serum samples. Pearson correlation coefficient (r = 0·868) demonstrates strong positive correlation between methods, indicating good agreement in sample ranking despite systematic differences in absolute values. *(B) Bland-Altman Plot*. Agreement analysis plotting the difference between methods against the average of both methods for each sample. *(C) Distribution Comparison*. Box plots comparing the distribution of NT50 values between eGFP and immunostaining methods across all 40 samples. *(D) Log-Scale Correlation*. Scatter plot showing the relationship between average eGFP NT50 values and immunostaining NT50 values for 40 serum samples with logarithmic scales. *(E) Variability in eGFP Replicates*. Histogram showing the distribution of coefficient of variation (CV) values for the three eGFP technical replicates across all samples. CV is calculated as (standard deviation / mean) × 100% and represents the intra-assay variability for each sample. *(F) Ratio Distribution*. Histogram displaying the distribution of ratios (eGFP / Immunostaining) across all samples.

Paired analysis revealed a systematic difference in absolute NT_50_ values between the two read-out methods (Wilcoxon signed-rank test, p < 0·001) (Figure 4B). Despite this shift, both read-outs consistently discriminated EBOV-positive and negative sera and preserved relative ranking across samples.

These findings demonstrate that fluorescence-based read-out provides a reliable and efficient alternative to immunostaining while retaining flexibility for laboratories with differing technical capabilities.

## Discussion

Neutralisation assays are a cornerstone of EBOV immunogenicity assessment and play a central role in vaccine and monoclonal antibody clinical trials, serosurveillance, and outbreak preparedness^8,15,29,33^. However, the lack of harmonisation across available neutralisation platforms complicates the interpretation and comparability of immunogenicity data generated in different studies and settings. In this study, we present a standardised microneutralisation assay based on biologically contained EBOV lacking the essential VP30 gene (EBOVΔVP30) and demonstrate its strong concordance with wild-type virus neutralisation while maintaining compatibility with BSL-2 laboratories.

A key finding of our work is the strong correlation between EBOVΔVP30 neutralisation titres and those obtained using live EBOV (Spearman ρ = 0·8725), exceeding correlations previously reported for pseudotyped virus systems^19^. This observation underscores the biological relevance of the EBOVΔVP30 platform and highlights important limitations of commonly used pseudotyped assays. While pseudoviruses provide valuable advantages in terms of safety and throughput, their reliance on heterologous viral backbones and single-cycle entry events may fail to fully capture the structural, genetic, and replicative complexity of wild-type filoviruses. Differences in virion morphology, glycoprotein processing, and co-transcriptional editing have been shown to influence antibody sensitivity and neutralisation profiles, potentially resulting in assay-dependent ranking of sera or therapeutic antibodies^19,23–25^. In the context of clinical trials, such variability may translate into inconsistent immunogenicity read-outs across vaccine platforms or study sites, complicating cross-trial comparisons and meta-analyses. Neutralising antibody titres are widely used as functional immunogenicity endpoints in EBOV clinical trials, although the precise correlates of protection remain incompletely defined.

By contrast, biologically contained EBOV systems preserve wild-type viral genome organisation, multi-cycle replication, and virion assembly while substantially reducing biosafety constraints^26–28^. Our data demonstrate that EBOVΔVP30 combines these features with robust discrimination between EBOV-positive and negative serum samples and strong alignment with live virus neutralisation outcomes. Importantly, the assay reliably reproduced borderline or low-titre responses previously observed in wild-type virus assays, suggesting that its performance reflects true biological variation rather than platform-specific artefacts. These findings position EBOVΔVP30 as a practical bridge between high-containment reference assays and lower-complexity surrogate systems.

Assay standardisation and scalability are critical considerations for clinical development and outbreak response. Live virus neutralisation assays, while biologically relevant, are poorly suited to high-throughput workflows, GCLP compliance, and decentralised testing due to their reliance on BSL-4 infrastructure and prolonged assay duration^17^. In contrast, the EBOVΔVP30 microneutralisation assay can be performed under BSL-2 conditions, supports multi-well plate formats, and provides a quantitative read-out within 45 hours. The use of the WHO International Standard for anti-EBOV immunoglobulin further facilitates calibration and inter-laboratory comparability, which are essential prerequisites for meaningful interpretation of immunogenicity endpoints in multicentre trials^30^.

An additional strength of the EBOVΔVP30 platform is the flexibility of its read-out. We show that fluorescence-based detection of eGFP-positive foci provides neutralisation titres that are highly concordant with those obtained using conventional immunostaining approaches. Although eGFP-based and immunostaining read-outs showed a systematic difference in absolute neutralisation titres, both methods were strongly correlated and reliably classified EBOV-positive and negative samples. This indicates that the observed difference reflects inherent read-out characteristics rather than reduced assay performance, supporting the use of eGFP fluorescence as an efficient and scalable alternative, provided that a single read-out strategy is applied consistently. Such adaptability is particularly relevant for laboratories in outbreak-prone or resource-limited regions, where infrastructure constraints may otherwise limit participation in global clinical research efforts.

From a translational perspective, our findings have direct implications for EBOV vaccine and monoclonal antibody development. Neutralisation assays are frequently used to rank candidate vaccines, assess dose–response relationships, and evaluate immune durability. Assay-dependent variability in neutralisation read-outs therefore has the potential to influence downstream clinical decision-making. By providing a biologically authentic yet accessible alternative to live virus assays, EBOVΔVP30 offers a platform that should improve the consistency and interpretability of neutralising antibody measurements across studies, provided that a single read-out strategy is applied consistently. This is particularly relevant for large, multi-site clinical trials and for initiatives aimed at assay harmonisation and regulatory alignment.

Beyond its immediate application in EBOV research, the successful implementation of a biologically contained EBOV neutralisation assay highlights the broader potential of this approach for other high-consequence pathogens. Similar systems have already been developed for Marburg virus and Sudan virus and have proven valuable for antiviral screening and mechanistic studies^26–28^. Extending biologically contained platforms to functional serology could substantially expand global capacity for immunogenicity assessment without compromising biosafety. Consistent with previous reports using biologically contained filoviruses, no evidence of VP30 reversion or loss of biological containment was observed, supporting the safety of this platform. Such advances would support outbreak preparedness, accelerate clinical development pipelines, and enhance the comparability of immune data generated across diverse settings.

In conclusion, we demonstrate that a biologically contained EBOVΔVP30 microneutralisation assay provides a clinically relevant, scalable, and standardisable alternative to existing neutralisation platforms. By closely mirroring live virus neutralisation while remaining compatible with BSL-2 laboratories and GCLP-aligned workflows, this assay addresses key limitations that currently hinder cross-trial comparability and assay harmonisation. Our findings support the integration of biologically contained EBOV systems into translational research and clinical development frameworks and provide a foundation for similar approaches targeting other emerging and re-emerging viral threats.

## Data Availability

All data produced in the present study are available upon reasonable request to the authors

## Contributors

JV, OD, DN, JAB, and SL performed experiments. JV, OD, DN, JAB, SL, PM, and MWC performed analysis. JAB provided clinical samples. PM and MWC conceptualised and supervised the project. PM and MWC verified the data and had full access to all the raw data in the study. All authors contributed to the writing of the manuscript. All authors read and approved the final version of the manuscript.

## Acknowledgments

MC received funds from Coalition of Epidemic Preparedness Innovations (CEPI) grant number H5R01920. We thank dr. Francesca Donnellan and dr. Pramila Rijal from Jenner Institute, University of Oxford, for kindly providing the human anti–Ebola virus glycoprotein monoclonal antibody used in this study.

