## Supplementary figures and images for "Biologically contained Ebola virus enables standardised neutralisation testing for preclinical and clinical immunogenicity assessment"

### supplementary Figure 1

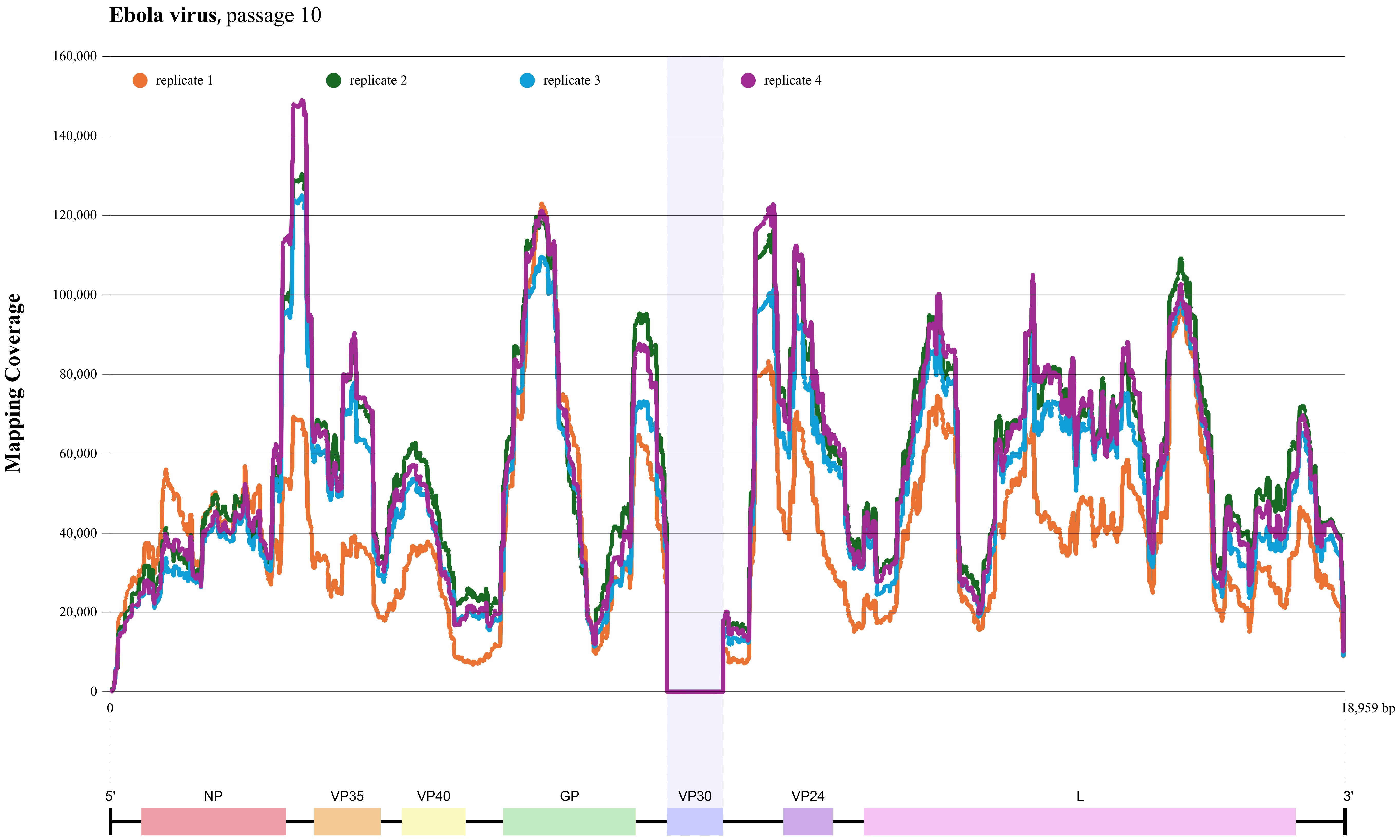
